# The Lasting Legacy of COVID-19: Exploring the Long-Term Effects of Infection, Disease Severity, and Vaccination on Health and Cognitive Function

**DOI:** 10.1101/2023.04.12.23288455

**Authors:** Jaroslav Flegr, Ashkan Latifi

**Affiliations:** Laboratory of Evolutionary Biology, Department of Philosophy and History of Sciences, Faculty of Science, Charles University; Viničná 7, 128 00, Prague, Czechia; Department of Psychology, Faculty of Economics and Social Science, Bu-Ali Sina University, Hamadan, Iran

## Abstract

COVID-19 affects a variety of organs and systems of the body including the central nervous system. Recent research has shown that COVID-19 survivors often experience neurological and psychological complications that can last for months after infection. We conducted a large internet study using online tests to analyze the effects of SARS-CoV-2 infection, COVID-19 severity, and vaccination on health, intelligence, memory, and information processing precision and speed in a cohort of 4,446 subjects. We found that both SARS-CoV-2 infection and COVID-19 severity were associated with negative impacts on patients’ health. Furthermore, we observed a negative association between COVID-19 severity and cognitive performance. Younger participants had a higher likelihood of SARS-CoV-2 contraction, while the elderly had a higher likelihood of severe COVID-19 and vaccination. The association between age and COVID-19 severity was primarily mediated by older participants’ impaired long-term health. Vaccination was positively associated with intelligence and the precision of information processing. However, the positive association between vaccination and intelligence was likely mediated by achieved education, which was itself strongly associated with the likelihood of being vaccinated.

## Introduction

SARS-CoV-2 appeared in Wuhan, China in December 2019. Although this viral infection was initially deemed to be a respiratory disease, researchers have since studied and discovered other aspects of its pathogenicity. For example, it is argued that COVID-19 is also a neuroinvasive disease that, by triggering a cytokine storm, can affect the nervous system and may potentially be involved in the onset and progression of neuropsychiatric and neurodegenerative complications ^1^. More specifically, three main pathophysiological manifestations of COVID-19 central nervous system involvement are: 1) thrombosis, 2) inflammation, and 3) direct damage to the CNS cells (see Manta et al, 2022, #27860). However, this review article suggests that the third possibility is less likely as there is no evidence that the virus has tropism in cells of the CNS or is capable of crossing the blood-brain barrier. Another recent review also proposes that, as evidence from molecular studies demonstrates, SARS-CoV-2 viral invasion of the brain is not a likely neuropathogenic mechanism (see Silva et al., 2022, #27861). As to the Blood-brain and/or blood-CSF barrier crossing of the virus, these researchers conclude it is more likely that SARS-CoV-2 spike protein crossing is the mechanism involved. It is possible that the primary pathophysiological mechanism is brain inflammation. As this review article maintains, infection in general is capable of affecting the function, morphology, and molecular biology of microglia, astrocytes, monocytes, and T-lymphocytes so that, as an effect of infection, these cells’ activity is upregulated resulting in aberrant production of cytokines such as tumor necrosis factor (TNFα), interleukin-1α, and interleukin-6 which in turn is known to play a role in brain/mental dysfunction.

A comprehensive review of the neuropsychiatric pathogenicity of long COVID reported that common complications included fatigue, cognitive impairment, sleep disorders, depression, anxiety, and post-traumatic stress disorder ^2^. A recent meta-analysis of a large dataset of 1.2 million individuals who had symptomatic COVID-19 also revealed that 6.2% of the subjects experienced 1 to 3 symptoms of long COVID, namely fatigue, mood swings, or bodily pain (3.2%, 95% CI, 0.6%-10%), cognitive problems after adjusting for health status before contracting COVID-19 (2.2%, 95% CI, 0.3%-7.6%), and ongoing respiratory problems (3.7%, 95% CI, 0.9%-9.6%). Models in this study estimated these to comprise 51.0% (95% CI, 16.9%-92.4%), 35.4% (95% CI, 9.4%-75.1%), and 60.4% (95% CI, 18.9%-89.1%) of long COVID cases, respectively ^3^.

Neurological sequelae such as brain fog, myalgias, headache, cognitive impairment, sensorimotor deficits, smell and taste disorders, mood disorders, sleep disorders, and fatigue in long COVID patients have been reported to be related to oxidative and neuroinflammatory processes caused by COVID ^4^. In this regard, abnormally elevated levels of TNFα have been proposed as the underlying cause of long-term impairment in executive functions in COVID-19 survivors even 15 months after discharge from the hospital ^5^. In addition to these, COVID-related hypoxia, COVID-19’s entrance into the central nervous system through the blood-brain barrier, and coagulation dysfunction have also been suggested as possible mechanisms of COVID-19-induced injury to the CNS ^6^.

A substantial body of research, including ^7^, has established a connection between impairment of cognitive functions and brain inflammation as also discussed earlier in our article. Accordingly, among other concerns, researchers have begun to investigate the possible COVID-induced cognitive impairment in individuals who contracted the disease. In a study using iPad-based online neuropsychological tests, researchers found that 29 patients who had recovered from COVID-19 did not differ from their matched controls in the Trail Making Test (assessing cognitive processing speed and executive functions), Sign Coding Test (investigating visual scanning and perception, memory, and eye movement), and Digital Span Test (testing concentration, resistance to information interference, and instantaneous memory). However, they differed in the Continuous Performance Test (measuring impulse and continuous and selective attention through a recognition task of a pair, two consecutive pairs, and three consecutive pairs of animal pictures in parts one, two, and three of the test, respectively), showing that patients who had COVID-19 demonstrated a lower reaction time in the first part (774.59 ± 119.33 vs 843.22 ± 140.97, p=0.050) and second part (817.06 ± 114.53 vs 879.59 ± 123.87, p=0.051), lower correct number of the second part (7.07 ± 2.45 vs 8.72 ± 1.79, p=0.050) and third part (6.34 ± 2.50 vs 8.21 ± 1.90, p<0.01), and higher missing number of the second (41.55 ± 2.90 vs 39.59, p<0.01) and third part (40.38 ± 3.10 vs 38.45 ± 2.13, p<0.01) of this test 27656. Their findings showed that sustained attention was significantly more compromised in the patient group than the control group. However, researchers investigating cognitive functioning in 54 patients who had mild COVID-19 and 36 matched healthy controls, using the Stroop test and Visual Aural Digit Span Test Form B (VADS-B), found that the patient group compared to controls had significantly higher Stroop test word reading-spontaneous correction number (0.13 ± 0.39 vs 0.00 ± 0.00, p<0.05) and reading time (19.1 ± 3.7 vs 16.3 ± 2.7, p<0.01); saying the word color - number of wrong words (0.46 ± 1.13 vs 0.03 ± 0.17, p<0.05), spontaneous correction number (0.78 ± 0.92 vs 0.33 ± 0.68, p<0.01) and saying time (36.2 ± 10.8 vs 28.5 ± 5.0, p<0.01); saying the box’s color - spontaneous correction number (0.2 ± 0.4 vs 0.0 ± 0.0, p<0.0) and saying time (12.1 ± 2.2 vs 10.2 ± 1.3, p<0.01), Stroop interference (11.7 ± 6.3 vs 8.0 ± 3.8, p<0.05) and speed factor duration (9.3 ± 1.5 vs 7.9 ± 1.2, p<0.01) collectively indicating worse attention, precision, and slower information processing in the patient group than the control group; in addition, the VADS-B test mean-scores were significantly lower in the patient group than in the controls (p<0.05); hence, according to these results, a decline in attention and short-term memory was observed in the patient group compared with the controls 27675 ^8^.

Regarding broader aspects of cognition, a study with a sample of 32 males and 182 females with a history of COVID-19 implementing tests for processing speed, attention, verbal, visual, prospective and working memory, executive functions, visuospatial skills, and language showed that more than 85% of the subjects had alterations in at least one of the tests. Attention displayed the largest deficit regardless of age; however, age moderated the decline in processing speed, verbal memory, executive functions, visual memory, and working memory. The authors reported that the declines were less pronounced in the elderly than the young, which they suggested to be due to a weakened immune system in the former; hence, a weaker autoimmune response and decreased inflammation in seniors ^9^. Another study used a comprehensive collection of tests, namely, MoCA and Clock-Drawing Test (global cognitive skills), Stroop test, Digit Span Test, and Trail Making Test (attention), Öktem Verbal Memory Processes Test (memory), Fluency Test (executive functions), Rey Osterrieth Complex Figure Test (visual perceptual skills), and Neuropsychiatric Inventory (neuropsychiatric status), to assess cognitive functioning and neuropsychiatric status in 50 subjects infected with COVID in the last 60 days and 50 non-infected subjects. The authors reported that global cognitive functions, memory functions, visual perceptual functions, and neuropsychiatric status, but not attention functions and executive functions (except for Phonemic fluency and TMT-A), were negatively affected by COVID-19 in the patients when compared with controls (p<0.05) ^10^. A longitudinal study (before and after the start of the pandemic) investigating cognitive decline in middle-aged and older adults with and without a history of mild COVID (52 and 41, respectively) using MoCA (Montreal Cognitive Assessments) also found that the MoCA mean-score was significantly lower in participants with a history of mild COVID than in controls (21.7 ± 4 vs. 19.6 ± 4.2, p<0.05). Additionally, multivariate logistic regression analysis showed that the odds ratio of experiencing cognitive decline was 18.1 times higher in the former group (95% CI 1.75–188, p<0.05) ^11^. Regarding COVID-induced cognitive impairments, it is argued that an inflammatory parainfectious process may be involved, which targets the frontal lobes or networks related to them. This is suggested by evidence of behavioral and dysexecutive symptoms observed in patients and neuropsychological findings pointing to frontal hypometabolism and fronto-temporal hypoperfusion in COVID patients ^12^.

As the aforementioned studies suggest, COVID-19 can negatively affect cognitive functions and health in patients with a history of acute and those with a history of mild COVID. However, most of these studies were based on small sample sizes and had somewhat controversial findings. For example, although Zhou et al. ^13^ found no significant difference between the patient group and controls in cognitive processing, executive functions, memory, concentration, and resistance to information interference, and they observed lower reaction times in the patient group, Demir et al. ^8^ reported higher reaction times, slower processing speed, and lower resistance to Stroop interference in their patient group compared with controls. Similarly, among other findings, processing speed, attention, working memory, and executive functions were found to be more compromised in patients in another study ^9^; however, other research did not find a significant difference between the patients and controls in attention and executive functions ^10^. The role of age has also proved controversial, as one study reported a negative association between age and impairment in cognitive functioning ^9^, while another study found a positive association in the patient group ^14^. Findings regarding COVID severity have also been somewhat controversial, as one study found no significant relationship between disease severity and cognitive deficits ^10^, while other studies reported significant associations ^15^. Furthermore, the possible interrelationships of age, long-term sickness, current sickness, and COVID severity with cognition have not been addressed in previous research.

With the objective of addressing these gaps in knowledge, the current cross-sectional study sought to investigate the impact of SARS-CoV-2 infection, COVID-19 severity, and COVID-19 vaccination on the cognitive performance and health outcomes of a cohort of 4,446 participants. We measured intelligence, memory, reaction times, and information processing speed using a set of four online tests, and then analyzed the data using partial Kendall Tau correlation tests and path analysis techniques.

## Results

The initial dataset comprised 5,230 participants who answered a question about their COVID status (66% women and 33% men). Out of these, 1,793 had not yet been infected, 2,246 had been diagnosed with COVID, 763 were possibly infected but not diagnosed, 22 were awaiting test results, and 406 had not been diagnosed but were in quarantine. Those who answered “No” or “No, but I was in quarantine” were categorised as COVID-negative (coded 0), while those who answered “Yes, I was diagnosed with COVID” were categorised as COVID-positive (coded 1).

Data from other subjects were excluded from the dataset (785 participants). Therefore, the final dataset included 2,199 COVID-negative and 2,246 COVID-positive participants (4,445 participants in total). Table 1 and Figures 1-3 present the descriptive statistics for the dataset.

**Table 1.**
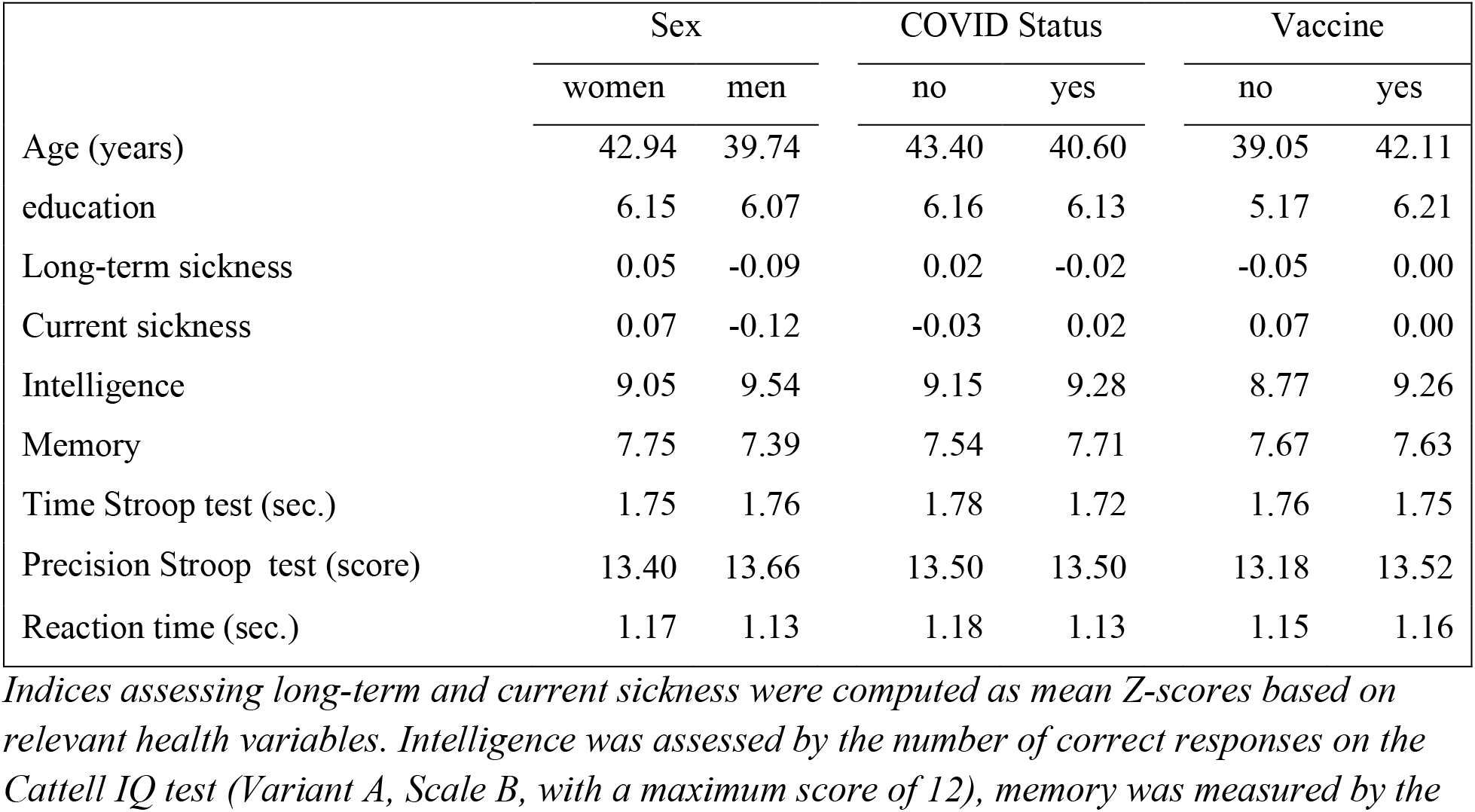
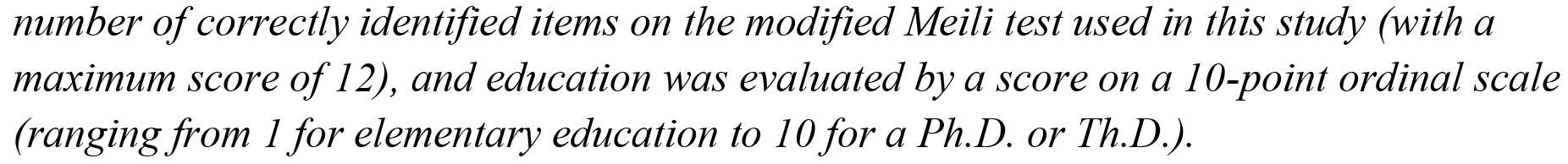
Descriptive statistics of the final set

|  | Sex |  | COVID Status |  | Vaccine |  |
| --- | --- | --- | --- | --- | --- | --- |
|  | women | men | no | yes | no | yes |
| Age (years) | 42.94 | 39.74 | 43.40 | 40.60 | 39.05 | 42.11 |
| education | 6.15 | 6.07 | 6.16 | 6.13 | 5.17 | 6.21 |
| Long-term sickness | 0.05 | -0.09 | 0.02 | -0.02 | -0.05 | 0.00 |
| Current sickness | 0.07 | -0.12 | -0.03 | 0.02 | 0.07 | 0.00 |
| Intelligence | 9.05 | 9.54 | 9.15 | 9.28 | 8.77 | 9.26 |
| Memory | 7.75 | 7.39 | 7.54 | 7.71 | 7.67 | 7.63 |
| Time Stroop test (sec.) | 1.75 | 1.76 | 1.78 | 1.72 | 1.76 | 1.75 |
| Precision Stroop test (score) | 13.40 | 13.66 | 13.50 | 13.50 | 13.18 | 13.52 |
| Reaction time (sec.) | 1.17 | 1.13 | 1.18 | 1.13 | 1.15 | 1.16 |

*Indices assessing long-term and current sickness were computed as mean Z-scores based on relevant health variables. Intelligence was assessed by the number of correct responses on the Cattell IQ test (Variant A, Scale B, with a maximum score of 12), memory was measured by the*
number of correctly identified items on the modified Meili test used in this study (with a maximum score of 12), and education was evaluated by a score on a 10-point ordinal scale (ranging from 1 for elementary education to 10 for a Ph.D. or Th.D.).

**Fig. 1.**
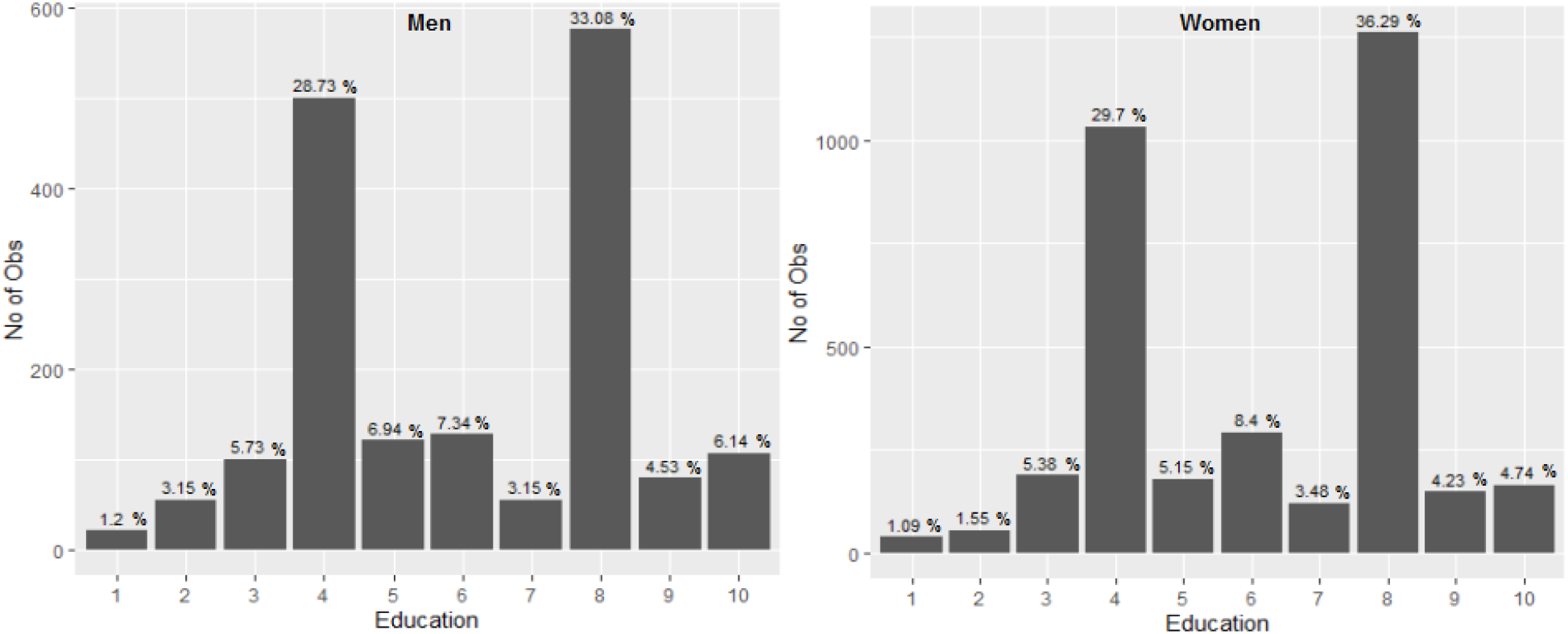
Education of female and male participants of the study *The codes 4 and 8 correspond to Complete secondary education or higher vocational training (A-levels or diploma), and master’s degree, respectively*.

**Fig. 2.**
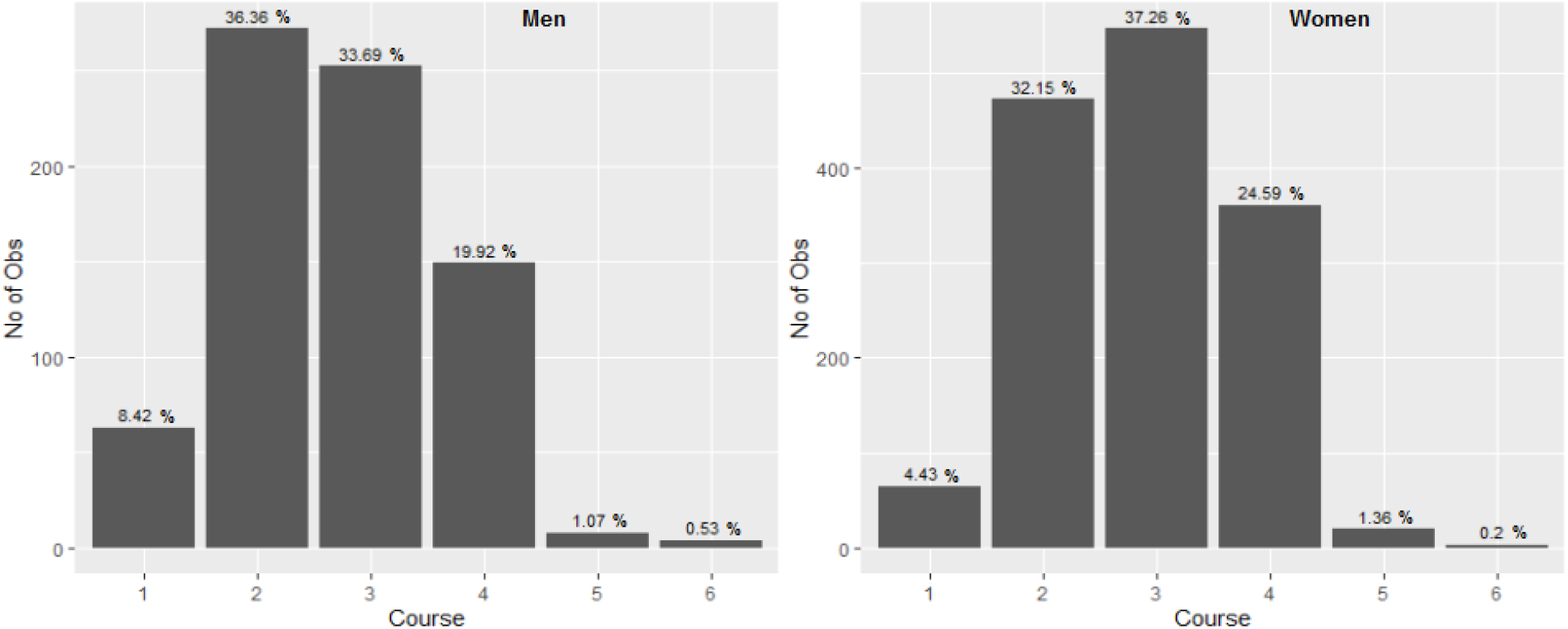
Severity of the course of COVID-19 in the female and male participants of the study *Codes 1 to 6 correspond to “No symptoms”, “Like mild flu”, “Like normal flu”, “Like severe flu”, “hospitalized”, and “at ICU”, respectively*.

**Fig. 3.**
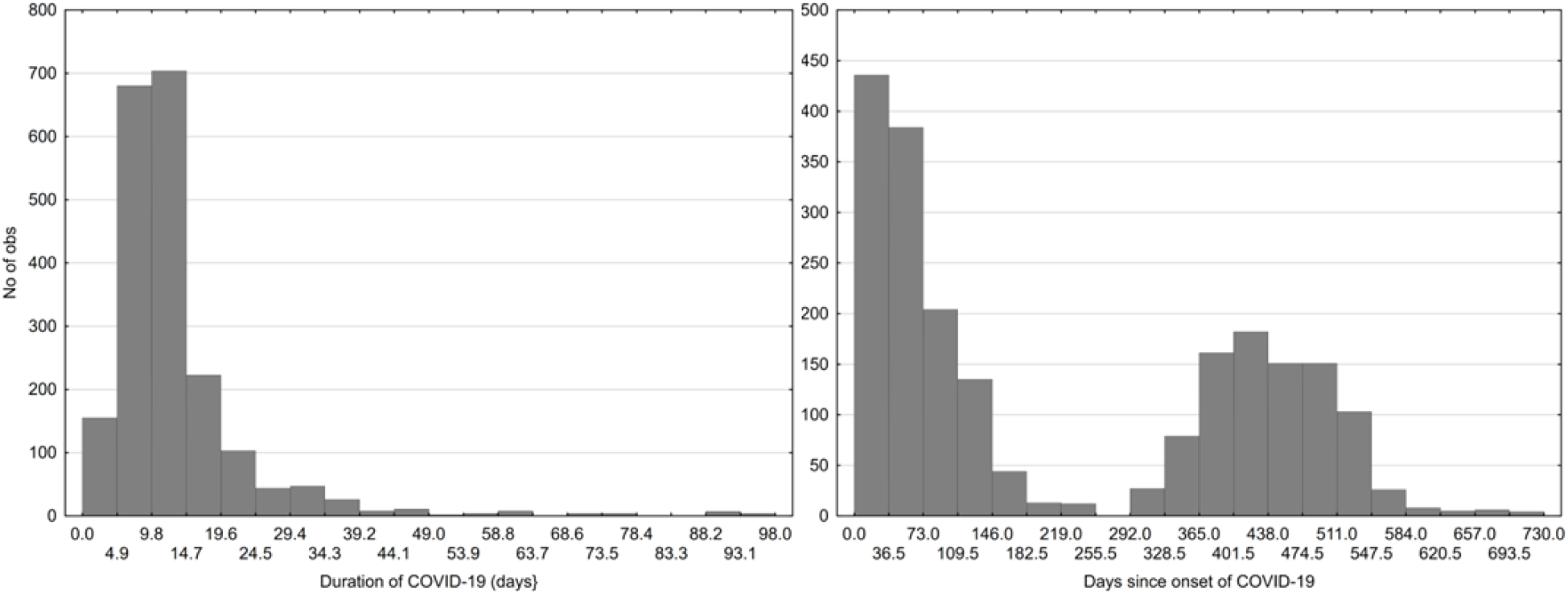
Duration of COVID-19 and time passed since start of the covid *The left histogram does not show 21 (1.02%) of the participants, including 17 (1.26%) women and 4 (0.56%) men, who reported a duration of COVID longer than 100 days*.

The study’s findings revealed that a higher percentage of men (52.32 %) were diagnosed with COVID compared to women (49.66 %), but this difference did not reach statistical significance (OR = 1.11, CI_95_: 0.97-1.26, p = 0.0969). Additionally, a significantly higher proportion of men (93.95%) reported being vaccinated compared to women (90.71%) (OR = 1.58, CI_95_: 1.26-2.01, p = 6.042E-05). The study also demonstrated that vaccinated individuals were less likely to have been diagnosed with COVID (49.05%) compared to those who were not vaccinated (67.24%), with a highly significant difference (OR = 0.469, CI_95_: 0.36-0.59, p = 7.366E-11). Although vaccinated subjects had a lower risk of hospitalization (1.21% vs. 1.72%) and treatment in an Intensive Care Unit (0.25% vs. 0.86%), the effect of vaccination on the course of the disease was not significant, likely due to the small number of non-vaccinated subjects and the low number of subjects with a severe course of COVID (Table 2) (correlation between vaccination and course of the disease, Spearman R = -0.01, p = 0.64).

**Table 2.** Course of COVID reported by vaccinated and non-vaccinated participants

|  | Vaccinated | No<br>symptoms | Mild flu | Normal<br>flu | Severe<br>flu | Hospital | ICU |
| --- | --- | --- | --- | --- | --- | --- | --- |
| Count | No | 11 | 73 | 96 | 47 | 4 | 2 |
| Percent |  | 4.72% | 31.33% | 41.20% | 20.17% | 1.72% | 0.86% |
| Count | Yes | 117 | 670 | 703 | 462 | 24 | 5 |
| Percent |  | 5.91% | 33.82% | 35.49% | 23.32% | 1.21% | 0.25% |
| Count | All | 128 | 743 | 799 | 509 | 28 | 7 |
| Percent |  | 5.78% | 33.56% | 36.09% | 22.99% | 1.26% | 0.32% |

**Table 3.** Correlation between COVID-related variables with age, sex, health, and performance

|  | Partial Kendall Tau |  |  |  |  |
| --- | --- | --- | --- | --- | --- |
|  | Infected | Course | Duration of COVID | Since COVID | Vaccinated |
| Age | **-0.080** | **0.051** | **0.097** | -0.007 | **0.060** |
| Sex | 0.017 | **-0.074** | **-0.052** | **0.033** | **0.061** |
| Education | -0.005 | **-0.039** | **-0.042** | 0.003 | **0.111** |
| Long-term sickness | -0.017 | **0.148** | **0.118** | -0.002 | **0.031** |
| Current sickness | **0.023** | **0.122** | **0.097** | **-0.032** | -0.012 |
| Intelligence | 0.018 | **-0.035\*** | -0.020 | 0.014 | **0.064\*** |
| Memory | **0.020** | **-0.043\*** | -0.009 | -0.000 | 0.008 |
| Precision Stroop test | -0.000 | **-0.031\*** | -0.008 | 0.007 | **0.048\*** |
| Time Stroop test (sec.) | -0.009 | 0.022 | **0.031** | -0.018 | **-0.022\*** |
| Reaction time (sec.) | **-0.040\*** | 0.025* | 0.021 | -0.027 | -0.013 |
|  | p values |  |  |  |  |
| Age | 1.02E-15 | 0.000254 | 3.10E-11 | 0.614952 | 6.56E-11 |
| Sex | 0.088933 | 1.28E-04 | 0.000363 | 0.021335 | 2.84E-11 |
| Education | 0.578293 | 0.005089 | 0.00382 | 0.798982 | 2.83E-33 |
| Long-term sickness | 0.085133 | 1.18E-25 | 1.08E-15 | 0.839863 | 0.000699 |
| Current sickness | 0.017378 | 7.01E-18 | 3.32E-11 | 0.025363 | 0.178982 |
| Intelligence | 0.065577 | 0.012089 | 0.169031 | 0.323175 | 3.64E-12 |
| Memory | 0.042148 | 0.002362 | 0.525511 | 0.986622 | 0.373842 |
| Precision Stroop test | 0.962318 | 0.028728 | 0.544026 | 0.586288 | 1.95E-07 |
| Time Stroop test (sec.) | 0.323233 | 0.113122 | 0.030953 | 0.192413 | 0.015192 |
| Reaction time (sec.) | 5.30E-05 | 0.074332 | 0.145262 | 0.057141 | 0.141735 |

*Sex was a binary variable, with women as 0 and men coded as 1. Negative Tau values, for example, indicate that women reported a more severe COVID course than men. Infected and vaccinated were binary variables (No: 0, Yes: 1), and the course of COVID was rated on a 6-point scale (1: no symptoms, 6: treated at the Intensive Care Unit). The duration of COVID was measured in days from the start to the end of COVID, and the time since COVID was measured in days from the start of COVID to completing the internet tests. Significant effects are printed in bold. Asterisks indicate effects that remain significant after a correction for multiple (5) cognitive performance tests.*

The initial analysis using F tests and inspection of histograms indicated that the variances also differed between infected and non-infected subjects and between men and women. Given this and also because of the strong effects of age and sex on performance and health, a multivariate nonparametric test, a partial Kendall correlation test controlled for age and sex, was used to examine the associations between COVID infection, course of COVID, and vaccination with the performance and health of subjects (Tab. 3).

Correlation tests cannot establish causality or distinguish between direct and indirect effects. The association between COVID and education is unlikely to be a result of COVID impacting education. However, the connection between COVID and IQ is not as clear-cut. To address these uncertainties, we used path analysis. The findings from the analyses are presented in Figures 3-6. To simplify the information and prevent redundancy, we have combined the verbal presentation of the results from path analysis (PA) with their interpretation in the following section.

**Fig. 4.**
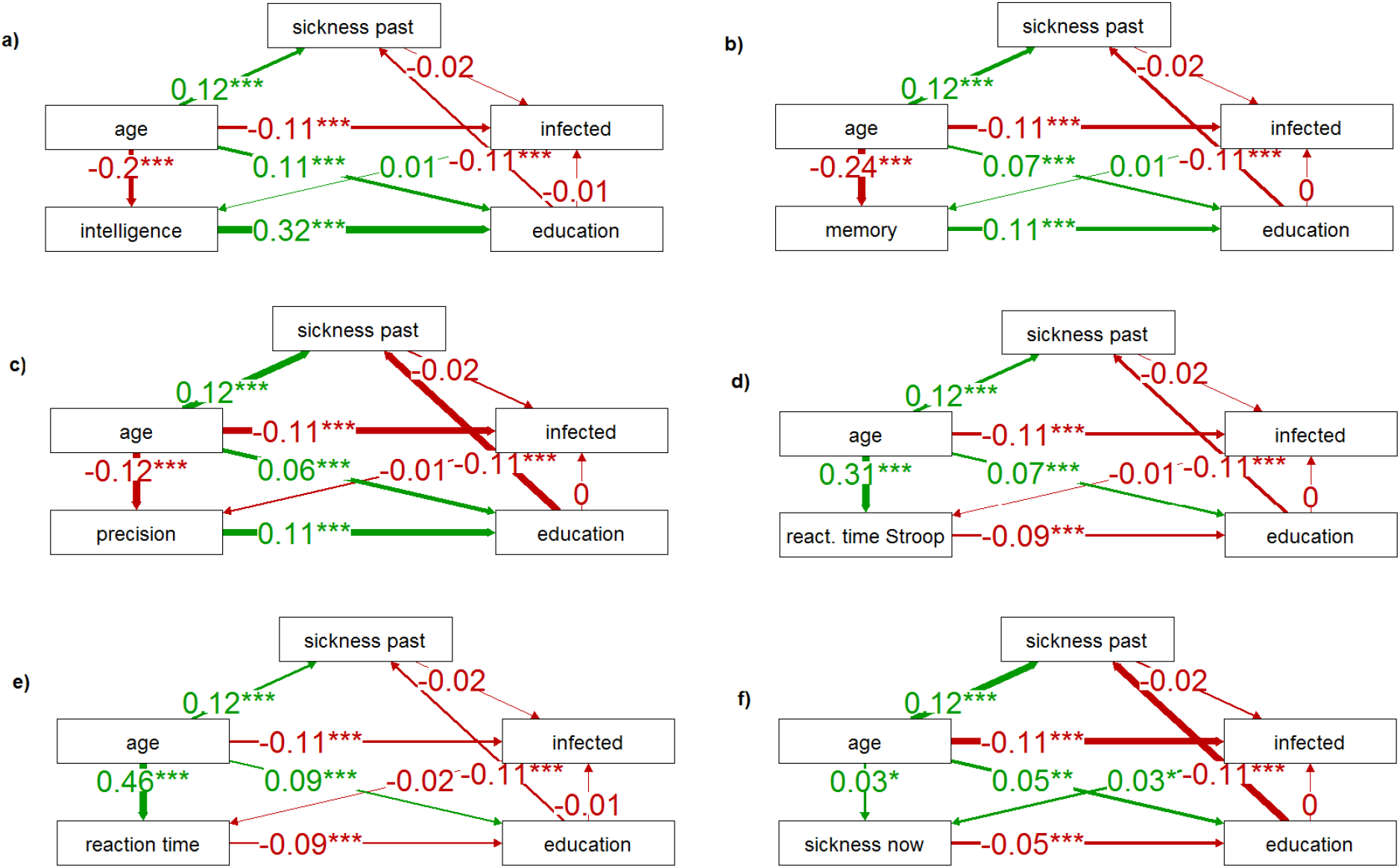
Correlation of being infected with SARS-CoV-2 with health and cognitive performance *Schemes a-e show the direct and indirect effects (path coefficients) of health and COVID-related variables on cognitive performance. The scheme f shows similar effects on the current sickness of the participants. The number of asterisks (one, two, or three) indicates their significance (0.05, 0.01, and 0.001, respectively). A positive path coefficient (green arrow) indicates that, for example, older participants reported a more severe COVID course than younger participants*.

**Fig. 5.**
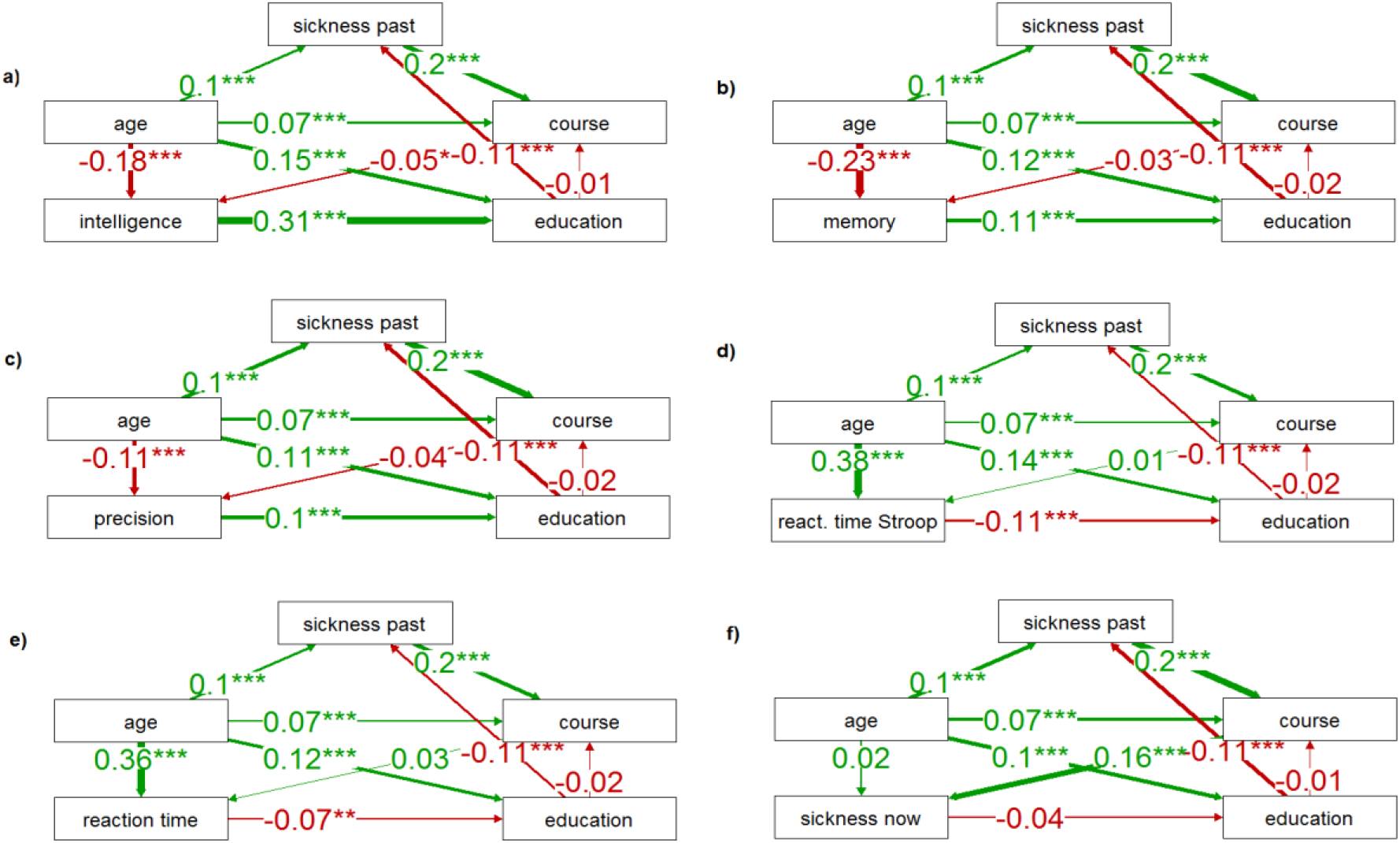
Correlation of COVID severity with health and cognitive performance *For legend see Fig. 4*

**Fig. 6.**
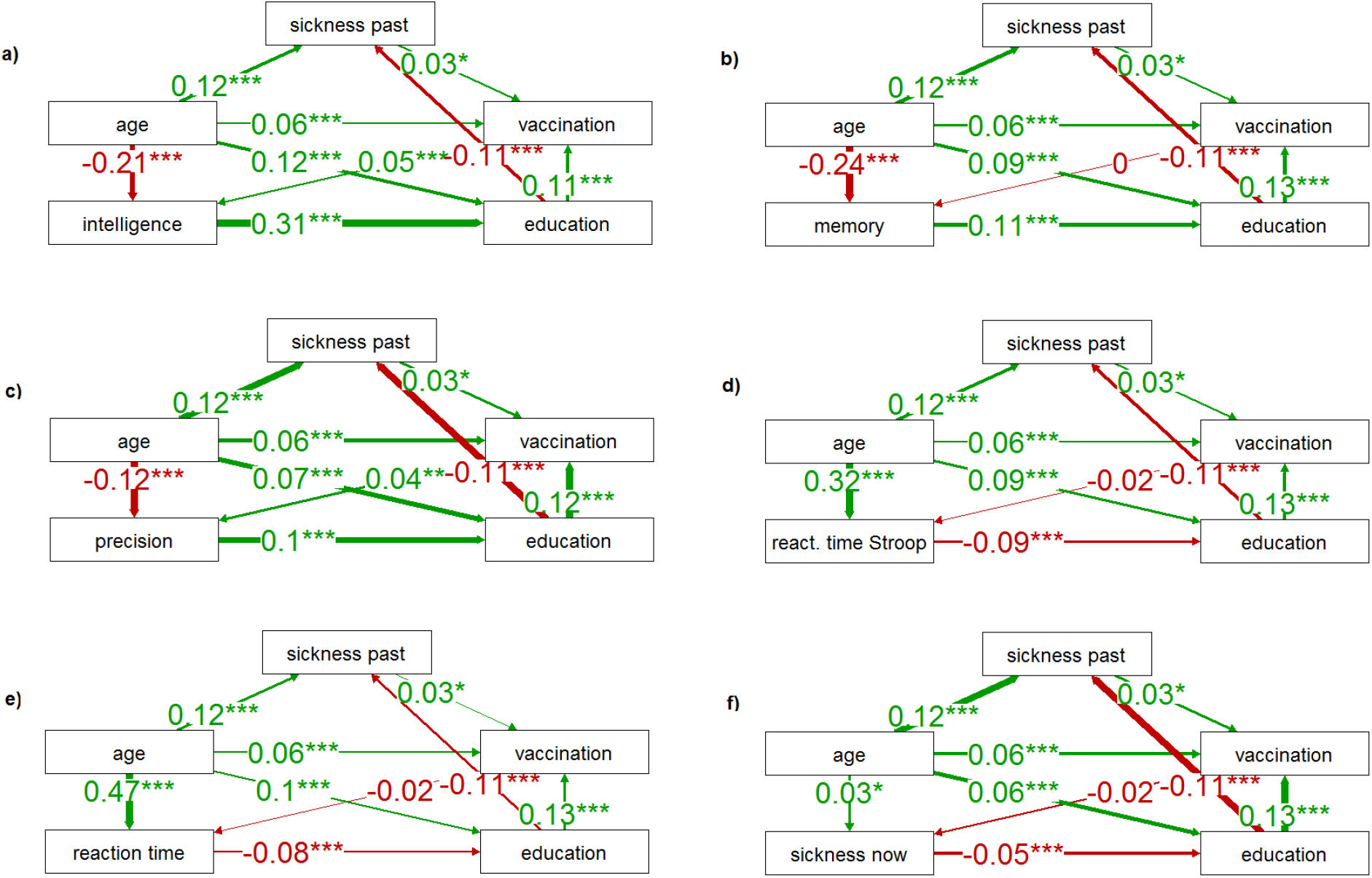
Correlation of vaccination and long-term sickness with current sickness and cognitive performance *For legend see Fig. 4*

The results of the PA for the binary variable infection (Fig. 4), specifically the comparison of path coefficients, indicated that the direct link between intelligence and infection was very weak and non-significant (0.01), but the positive impacts of intelligence (0.32), memory (0.11), and precision (0.11), and negative impacts of speed of information processing measured with the Stroop (−0.09) and simple reaction time tests (−0.09), and current sickness (−0.05) on achieved education were relatively strong and significant. Additionally, achieved education had a significant and negative impact on long-term sickness (−0.11), and long-term sickness may have had a slight negative effect on the risk of COVID infection (−0.02, not significant). This negative correlation may reflect the efforts of people with known health risks to avoid infection. The results also revealed that infection had a positive effect on current sickness (0.03). As to age, it had positive and significant effects on education (0.05-0.11), long-term sickness (0.12), reaction times (0.31-0.46), and current sickness (0.03) and significant and negative impacts on IQ (−0.2), memory (−0.24), precision (−0.12), and infection (−0.11).

Path analysis produced clearer results for the ordinal variable representing the severity of COVID (Fig. 5). The analyses showed once again that the positive influences of intelligence (0.31), memory (0.11), and precision (0.1), and the negative influences of the speed of information processing measured with the Stroop (−0.11) and simple reaction time tests (−0.07), and current sickness (−0.04, not significant) on achieved education were also relatively strong. Nevertheless, the analyses also revealed that long-term sickness increased the likelihood of a severe COVID course (0.2), and that a severe course was associated with worse cognitive test performance except for reaction times (IQ: -0.05, memory: -0.03, precision of information processing: -0.04). However, only the effect on IQ was significant (IQ: p = 0.036, memory: p = 0.126, precision: p = 0.100). A severe course of COVID-19 had a strong adverse effect on current sickness (0.16, p<0.01). These results indicated that the negative association between education and the course of COVID detected by partial Kendall tests was mediated by the negative effect of education on long-term sickness and the positive effect of long-term sickness on COVID severity. The effect of COVID severity on cognitive test performance as well as current sickness, was most likely direct. In addition, age had positive and significant effects on long-term sickness (0.1), course (0.07), education (0.10-0.15), reaction times (0.36-0.38), and current sickness (0.02, not significant though). The impact of age on the course was also indirectly mediated by long-term sickness suggesting that as the individual becomes older their history of health issues negatively affects the course of COVID-19 and makes it more likely for them to develop the severe form of the disease. Age again had a significant and negative impact on performance in cognitive tests (intelligence, -0.18; memory, -0.23; precision, -0.11; reaction times, 0.38 and 0.36).

The results of the path analysis for the binary variable vaccination are presented in Figure 6. The analysis showed that there was a significant positive correlation between poor health (Long-term sickness) and the likelihood of being vaccinated (0.03), supporting the idea that individuals who are aware of their health problems are more likely to try to avoid infection through vaccination. Additionally, higher levels of education were strongly correlated with a higher likelihood of being vaccinated (0.11-0.13). Being vaccinated also correlated positively with performance in cognitive tests, except for memory, but only the correlations with intelligence (0.05) and precision measured with the Stroop test (0.04) were significant. Moreover, being vaccinated was negatively correlated with current sickness (−0.02), but the effect was not significant (p = 0.190). It is possible that this correlation is due to the lower probability of a severe course of COVID in vaccinated individuals. The correlations of long-term sickness, vaccination, education, reaction times, current sickness, and performance in cognitive tests with age also conceptually yielded the same results as those reported in Figure 5.

## Discussion

Our cross-sectional study involved 2,246 volunteers who had contracted COVID-19 and 2,199 who had not. The study revealed that the infection itself had a weak but notable negative impact on the subject’s current health and a negligible effect on participants’ performance in cognitive tests. However, the severity of COVID-19 had a negative impact on current health that was approximately five times stronger than the sole infection and a significant negative effect on intelligence but not performance in other cognitive tests. On the other hand, vaccination had no effect on current health but significant and positive effects on performance in cognitive tests of intelligence and precision. Our findings also demonstrated that age had a positive and significant effect on being vaccinated and severity of COVID and a negative and significant impact on catching the disease. The positive effects of age on vaccination and COVID severity were also positively and significantly mediated by long-term sickness.

The negative impact of COVID-19 on current health found in our study was in agreement with the findings of the studies that reported similar adverse effects on COVID-19 patients’ current health ^16-20^. These negative effects even months after recovery may be the result of direct viral tissue damage afflicted by the virus on the host cells via its entry receptor ACE2, autoreactive T cells causing immune system dysregulation, SARS-CoV-2-specific CD8 T cell response, COVID-related cardiovascular and pulmonary complications, and/or neuroinflammation ^21,22^.

Our results also showed that the effects of COVID-19 infection on cognitive performance and functions were weak; a finding that was in disagreement with the former studies that reported significant adverse effects of both mild and acute COVID-19 on cognition ^8,9,11,23^. This finding was somewhat in agreement with Zhou et al. ^13^ who reported no significant difference between their patient group and controls in cognitive functions except for continuous and selective attention, and also another study that found no significant difference between the patient group and controls in terms of cognition ^24^. Many of the studies that reported impairment in cognitive performance and functions recruited subjects with a history of acute COVID (e.g., ^25,26^).

However, our dichotomous classification of the participants into either infected or non-infected groups may have resulted in losing the more specific impact of the severity of the disease on cognitive performance and function; hence, the non-significant results. However, this issue was addressed through our analyses of COVID severity in the current study. The analyses further revealed that being infected with COVID had positive effects on intelligence and memory and negative impacts on precision and reaction times; although these effects were non-significant and weak, these findings were in disagreement with those studies that reported impairment in cognitive functions in COVID patients ^8^ and in agreement with a study in which lower reaction times were reported in the patient group ^13^.

As to age, it was negatively and significantly associated with infection and cognitive functions (intelligence, memory, and precision) and positively and significantly with reaction times. The negative correlation between age and risk of contracting COVID suggests that the older adults were presumably more cautious of this disease because of their subjective perceived vulnerability to it, the news on higher rates of mortality and COVID complications in the elderly, or both. This indicates that behavioral immunity, i.e., the ability to avoid infection, was a relatively effective method of combating COVID-19, at least before the emergence of more contagious variants of the SARS-CoV-2 virus. These findings, regarding age and cognition, were in agreement with those studies that reported cognitive impairment in older adult COVID patients ^27-29^ and disagreement with a study in which the elderly were found to be less adversely affected by COVID than the young ^9^. However, in case of this latter study, the comparisons made between the three patient age groups (young, n= 29; middle age, n=97; and senior, n=88) were based on relatively disproportionate and small sample sizes that may have influenced their results. In addition, our findings demonstrated that a higher percentage of men (52.32%) compared with women (49.66%) were infected with COVID-19. Although the difference was not significant, this was in agreement with those studies in which men were found to be more vulnerable to COVID ^30^; ACE2 gene’s being located on the X chromosome is argued to be one of the principal reasons for this greater vulnerability to COVID in men ^31^.

Our analyses further suggested that COVID severity significantly negatively correlated with intelligence in the patient group. In addition, the performance measured as reaction times, precision, and memory were also negatively (but non-significantly) correlated with the severity of the disease; the negative effect of the severity of the disease on the performance in all five tests suggests that the total effect is statistically significant (p = 0.031, Fisher exact test). No previous study investigated the impact of COVID severity on patients’ intelligence; however, regarding processing speed, memory, and executive functions, our finding was in agreement with a study that reported no relationship between COVID severity and cognitive deficits ^10^ and in disagreement with those studies that reported a relationship ^15^. It may be that the very small fraction of patients with acute COVID in our sample resulted in these non-significant findings as these cognitive impairments are mostly found in patients with acute COVID who are reported to have higher levels of cytokines compared to mild COVID patients ^32^. It might be indicative that our results showed a non-significant positive correlation between performance in four of five performance tests – the fifth test, the memory test, showed no correlation (Tau 0.000). In this regard, the decline in cognitive functions could be physiologically related to the acute stage-related inflammation caused by SARS-CoV-2. Higher levels of TNF-alpha, IL-2R, IL-6, IL-10 are reported in COVID patients and it is found that there is an association between the attributed neuroinflammation and cognitive decline ^5,33,34^.

Regarding our findings, it may also be the case that intelligence as a more global cognitive component compared with other cognitive functions is more adversely affected by COVID; future research is needed in this regard. In addition, there was a strong negative impact of COVID severity on current health in our study which was in agreement with previous findings ^17,35^. This is also likely to be mainly an effect of COVID-induced inflammation in patients, see ^36^. Our results as well demonstrated that age had a significant and positive association with the course of COVID so that the older the patient, the more severe the disease. The results of our study align with previous research findings ^37,38^, which suggest that older adults may be more vulnerable to the effects of COVID-19 due to their weaker immune system ^39^. The results of our path analyses suggest that the association between age and COVID-19 severity is primarily mediated by long-term health issues, but age seems to have also a direct effect on the course of COVID. However, it is likely that older participants may have some health issues of which they are not aware and the existence of these issues can be responsible for the seemingly direct effect of age on the severity of the course of COVID.

Our research also revealed that there was a positive and significant association of vaccination with intelligence and precision. It could be hypothesized that vaccination may have resulted in reduced levels of stress of contracting COVID in individuals, which in turn could have culminated in their better cognitive performance compared with those unvaccinated who could have experienced higher levels of COVID-19 contraction stress, as there is evidence that demonstrates that lower levels of stress are associated with enhanced cognitive performance ^40^. Another explanation might be that vaccination is an effect rather than a cause in the observed association. Path analysis showed that intelligence had a strong positive effect on education and education in turn had a positive effect on the probability of being vaccinated. These two associations might have resulted in the observed relation between vaccination and intelligence. It must be reminded, however, that memory had also a strong effect on education despite no association between memory and vaccination was observed. It is, of course, possible that intelligence, but not memory, had also a direct effect on the probability of being vaccinated (standard path analysis cannot show the direction of arrows). Age was positively associated with vaccination, both directly (Tau = 0.06) and indirectly, through the mediation effect of long-term sickness. This finding suggests that older individuals were more likely to get vaccinated, possibly due to their higher perceived vulnerability to COVID, including the higher risk of mortality and serious COVID-related complications. Our study also found that older subjects had a lower probability of acquiring infection but a higher risk of a more severe course of COVID.

In addition, our results demonstrated that a significantly higher percentage of men (93.95%) compared to women (90.71%) were vaccinated against COVID. This may reflect the influence of the information about COVID-19 risk factors in the news on the public as men are found and reported to be more vulnerable to COVID-19 ^30^, this may have encouraged them to get vaccinated more readily. However, in the population under study, the men reported a less severe course of COVID than women. We also discovered that vaccinated individuals had a significantly lower likelihood of being diagnosed with COVID-19 (49.05%) compared to non-vaccinated ones (67.24%). However, it is highly likely that individuals who protect themselves against COVID-19 through vaccination also employ other protective measures such as mask-wearing and social distancing. Therefore, it is unclear what the proximal cause of the lower infection risk in vaccinated participants is.

### Strengths and Limitations

The study’s main strength was its large number of participants. In the majority of studies published so far, the number of participants has been considerably lower, or the cognitive performance of participants was assessed based on their subjective ratings rather than objective performance tests ^3^. Furthermore, our study used a natural cohort of internet users, which allowed for typical representations of subjects with different degrees of severity of COVID-19, rather than focusing solely on patients with severe cases as in many previous studies. An additional advantage of the current study was that participants were not informed in advance that one of the aims was to investigate the effects of COVID-19 and vaccination on health and performance. During recruitment and on the informed consent webpage, participants were told that the study would examine which biological and psychological factors influenced their performance test scores and moral attitudes. Questions about COVID-19 were placed towards the end of the 40-60 minute questionnaire, after all performance tests, to avoid any conscious or unconscious distortion of results resulting from participants’ subjective opinions about the positive and negative effects of COVID-19 or vaccination on health and cognition. (This procedure was approved in advance by the IRB). Another strength of the study was that participants were not given any rewards for their participation, which reduced the likelihood of “professional questionnaire fillers” or bots taking part.

The main limitation of the study is that the participants were self-recruited, and therefore, they do not represent a typical Czech population. Curious, altruistic people with interests in science were clearly overrepresented in the sample. Therefore, caution must be exercised when attempting to generalize the findings. Additionally, the physical health of the participants was self-reported and not examined by a medical professional. Although the study asked specific questions about the participants’ health and calculated health indices based on their responses, subjective factors such as hypochondria and overall psychological attitudes might have influenced the resulting indices.

The strength of the observed effects, which is the fraction of total variability in focal variables attributed to COVID or vaccination, may seem relatively low. For instance, the partial Kendall Tau of -0.042 for the effect of severe COVID on the participants’ performance in the memory test corresponds to Cohen’s f of 0.06, which is typically categorized as a small effect (but not negligible). Small effect sizes are typical for this type of study, as test performance can be influenced by several factors, including the motivation of the participants and the precision, reliability, and reproducibility of the tests.

## Conclusions

Our study reveals that the after-effects of COVID-19 persist for several months, even in individuals with mild symptoms cases. This underscores the importance of factoring in protection against the disease’s long-term negative impact on cognitive performance when evaluating the costs and benefits of preventive measures, such as vaccination. We found that, besides age, sex, and long-term health conditions, education plays a vital role in reducing the risk of SARS-CoV-2 infection and severe COVID-19. The positive effect of education is likely universal, also influencing the risk of other diseases and the likelihood of vaccination through its impact on long-term health. As a result, it is essential to implement initiatives aimed at enhancing the population’s education level as part of comprehensive strategies to protect public health in both developing and developed countries.

## Materials and Methods

### 1.1. Participants

An electronic survey consisting of several questionnaires and performance tests, with only some related to the present study, was advertised on Facebook and Twitter as a project “studying the interconnections between moral attitudes, cognitive performance, and various biological, psychological, and sociodemographic factors.” On the first page of the first questionnaire, participants were informed that the study was anonymous and that they could discontinue their participation at any time. They were also given the following information: “We will be determining which biological and psychological characteristics influence the results of performance tests and moral attitudes. We will measure your memory, speed, ability to concentrate, and intelligence.” Only those who confirmed they were over 15 years old and provided informed consent by clicking the corresponding button were allowed to participate in the study. The study was partially or fully completed by 8,800 subjects between March and June 2022. The project, including the method of obtaining informed consent, was approved by the Institutional Review Board of the Faculty of Science, Charles University (No. 2021/4).

### 1.2. Questionnaires and tests

In the survey, we assessed the intelligence of the subjects using the Cattel 16PF test (Variant A, Scale B) ^41^and their memory with a modified Meili test ^42,43^. During the Meili test, participants were first shown 12 words (knife, frog, pump, chain, tree, collar, ice, glasses, arrow, train, bars, rifle) for 24 seconds and then, about 30 minutes into the survey, they were asked to recall these words from a list of 24. Psychomotor performance (reaction time and precision) was measured using the Stroop test. This version of the Stroop test consisted of three parts with breaks for instructions and rest in between. In Part A, participants had to select a specific word (e.g. “red”) from a set of four words (“red,” “green,” “blue,” “brown”) displayed in a random order in the center of the screen. The words were written in a font color that did not match their meaning.

The command specifying which word to select was written in the upper part of the screen and participants were instructed to ignore the font color. In Part B, the stimuli were the same, but participants were asked to select a word written in a specific color, ignoring the meaning of the displayed words. Part C was similar to Part A, but the command specifying which word to select was always written in a different color that did not match either the meaning or color of the displayed stimuli. Before each part, participants received instructions on the rules, were informed of how many times the test would run (always five times), and were asked to react as quickly as possible. Participants could start each part of the Stroop test by pressing the “Start test” button. A similar test was utilized to measure reaction times. Participants were instructed to press a button corresponding to a specific character, which included the letters A, B, C, and D. The characters were presented in random order and all appeared in the same color (black). This variant of the test was administered eight times at the beginning of the experiment.

In the anamnestic part of the questionnaire, participants answered 11 questions about their long-term physical health. They had to respond to the questions about frequencies of doctor visits, fatigue, headaches, other physical pain, neurological diseases, and other chronic physical issues using 6-point ordinal scales. They also had to report the number of non-mental health medications prescribed by a doctor they were currently using (10 meant 9 or more), and the number of times they spent more than a week in the hospital in the past five years (5 meant 6 or more times). Participants were also asked about their usual physical feeling (5-point scale). This question was asked two times, once at the beginning of the questionnaire and once near its end, i.e. about 30 minutes later. Finally, they were asked how many years they expected to live (1: more than 99, 2: 90-99, 3: 80-89, 4: 70-79, 5: 60-69, 6: less than 60).

In addition to rating their typical physical state, participants also rated their current physical state on a 5-point scale at the beginning and towards the end of the questionnaire. Indices for long-term physical problems (long-term sickness) and current physical health issues (current sickness) were calculated as the mean Z-scores of the relevant questions ^44^.

In the anamnestic part of the survey, participants were also asked about their age, sex (men coded as 1, women coded as 0), education (ordinal scale 1–10: 1 – Basic education only, 2 – Basic education plus studying at secondary school, 3 – Secondary education including vocational training (without A-levels), 4 – Complete secondary education or higher vocational training (A-levels or diploma), 5 – Complete secondary education or higher vocational training, plus studying for a bachelor’s degree, 6 – Bachelor’s degree (BA, BSc), 7 – Studying for a master’s degree, 8 – Master’s degree (MA, MSc, MBA, MD, LL.M, MEng, etc.), 9 – Master’s degree, plus studying for a doctoral degree, 10 – Doctoral degree (PhD, DPhil, EdD, etc.)) and if they had been infected with SARS-CoV-2 (had COVID) (1: “not yet,” 2: “yes, I was diagnosed with COVID,” 3: “probably yes, but I was not diagnosed with COVID,” 4: “I am waiting for the result of a diagnostic test,” 5: “No but I was in quarantine”). For purposes of this study, answers 1 and 5 were coded as 0 (COVID-negative), answer 2 as 1 (COVID-positive), and answers 3 and 4 were coded as NA (data not available) as these participants’ COVID-19 infection history could not be reliably ascertained. If they answered “yes, I was diagnosed with COVID,” they were asked for the start and end dates of their COVID illness and to rate its severity on a 6-point scale (1: “No symptoms,” 2: “Like mild flu,” 3: “Like normal flu,” 4: “Like severe flu,” 5: “I was hospitalized,” and 6: “I was treated at an Intensive Care Unit”). The participants were also asked if they had been vaccinated against COVID (binary variables 0/1; we did not discriminate how many doses they received).

### 1.1. Data analyses

The influence of infection, COVID severity and duration, time since COVID onset, and vaccination status on health and cognitive performance were assessed using non-parametric partial Kendall correlation tests. These tests were run using the R script Explorer v. 1.0 ^45^, which utilizes the ppcor R package ^46^. In the analyses of the mixed sample of men and women, we controlled for both age and sex, while in the separate analyses for men and women, we controlled only for age. The Kendall correlation test allows for controlling confounding variables and is robust against the presence of outliers and the distribution shape of variables in general.

To account for multiple testing, we controlled the effects using the Benjamini-Hochberg procedure, with a false discovery rate (FDR) set at 0.10 ^47^. Path analysis was conducted using lavaan v. 0.6.12 ^48^ and semPlot 1.1.6 ^49^. The dataset is publicly available on Figshare ^50^.

## Data Availability

All data are available in the public repository figshare 10.6084/m9.figshare.14685825.v1

https://doi.org/10.6084/m9.figshare.22586446

## Technical notes

Throughout the article, the term “effect” is used in a statistical sense, meaning an observed association – the difference between the true population parameter and its null hypothesis value. Only in the Discussion section do we distinguish between cause and effect. Some parts of this study had a confirmatory, others an exploratory character. We, therefore, report the results corrected and non-corrected for multiple tests and discuss not only the formally significant effects but also trends that were not formally significant.

## Acknowledgment

This research was supported by Czech Science Foundation, grant number 22-20785S. Our sponsor had no involvement in the study design, the collection, analysis and interpretation of data, the writing of the report, or in the decision to submit the article for publication.

## Author contributions

JF planned the study and collected data, JF and AL analysed the data and wrote the article.

## Data availability

All data are available in the public repository figshare 10.6084/m9.figshare.14685825.v1

